# Variation at Spike position 142 in SARS-CoV-2 Delta genomes is a technical artifact caused by dropout of a sequencing amplicon

**DOI:** 10.1101/2021.10.14.21264847

**Authors:** Theo Sanderson, Jeffrey C. Barrett

## Abstract

Public SARS-CoV-2 genomes from the Delta lineage show complex and confusing patterns of mutations at Spike codon 142, and at another nearby position, Spike codon 95. It has been hypothesised that these represent recurrent mutations with interesting evolutionary dynamics, and that these mutations may affect viral load. Here we show that these patterns, and the relationship with viral load, are artifacts of sequencing difficulties in this region of the Delta genome caused by a deletion in the binding site for the 72_RIGHT primer of the ARTIC V3 schema. Spike G142D should be considered a lineage-defining mutation of Delta.

## Introduction

The ARTIC Network amplicon protocol is one of the most widely used approaches to sequence SARS-CoV-2 genomes. It consists of approximately 100 pairs of PCR primers which each amplify a ∼340 base-pair section of cDNA from the SARS-CoV-2 genome for subsequent sequencing. Version 3 of the ARTIC schema was designed using the Wuhan-Hu-1 reference sequence (MN908947) and released in March of 2020 (1, 2). Mutations that arise in SARS-CoV-2 in these primer sites might affect how well each section of the genome is amplified, and thus the quality and completeness of the resulting sequence. If such mutations increase in frequency, for example by occurring on widespread lineages, they can cause substantial issues of interpretation when using public sequence datasets.

Here we investigate the effect of a deletion found in the genome of all Delta lineage SARS-CoV-2 that profoundly reduces the amplification of ARTIC V3 amplicon 72. In particular we investigate its effects on calls for Spike mutations G142D and T95I, which have recently been argued to be recurring within the Delta lineage (3) and associated with higher viral load. We show that these observations are actually a direct consequence of amplicon 72 failure, and that G142D is a lineage-defining mutation for Delta.

## Results

Except where otherwise specified, the following analyses were performed on a set of COG-UK genomes (4) generated by the Wellcome Sanger Institute (5) from 1 March to 30 June 2021 for which *C*_*t*_ value (the PCR cycle threshold for detecting the virus in the diagnostic test – higher numbers mean less virus in the sample) data were available.

### In UK data, all Delta sequences with a known residue at Spike position 142 are G142D

A convenient, human-readable way to represent a SARS-CoV-2 genome is to simply list all positions that differ from the Wuhan reference, either in nucleotide or amino acid coordinates. This summary is available, for example, from GI-SAID in the `metadata.tsv` files. When represented this way, just 65% of our set of UK genomes are annotated as having the G142D mutation, and there is an unspoken assumption that the remaining 35% of sequences *do not* harbour the mutation. However, there is a third possibility: that we lack information on the residue at Spike position 142.

To distinguish these possibilities, it is necessary to look at the nucleotides corresponding to position 142 in a multiple-sequence alignment. We have built a simple tool to make converting from amino acid to nucleotide coordinates more straightforward (codon2nucleotide.theo.io, see also Table 1). Spike position 142 is encoded by a codon at nucleotide positions 21986-8. In the case of Delta, the relevant nucleotide mutation is a mutation at position 21987 from G in the reference sequence to A in Delta, which creates the Spike G142D substitution. Plotting the distribution of residues in our dataset shows that 65% of Delta sequences indeed have an A at the 21987 position – however nearly all of the remaining 35% of sequences do not have the G seen in the reference (and 100% of Alpha sequences), but instead have an N indicating that the nucleotide at this position is unknown (Fig. 1).

**Table 1.** Summary of the true nucleotides at each position discussed in this work in the Wuhan reference, Delta lineage, and AY.4 sublineage, and the amino acid mutations they result in.

| Genomic position | Reference nucleotide | Delta nucleotide (AA mutation) | AY.4 nucleotide (AA mutation) |
| --- | --- | --- | --- |
| 21987 | G | A (G142D) | A (G142D) |
| 21846 | C | C | T (T95I) |

**Fig. 1.**
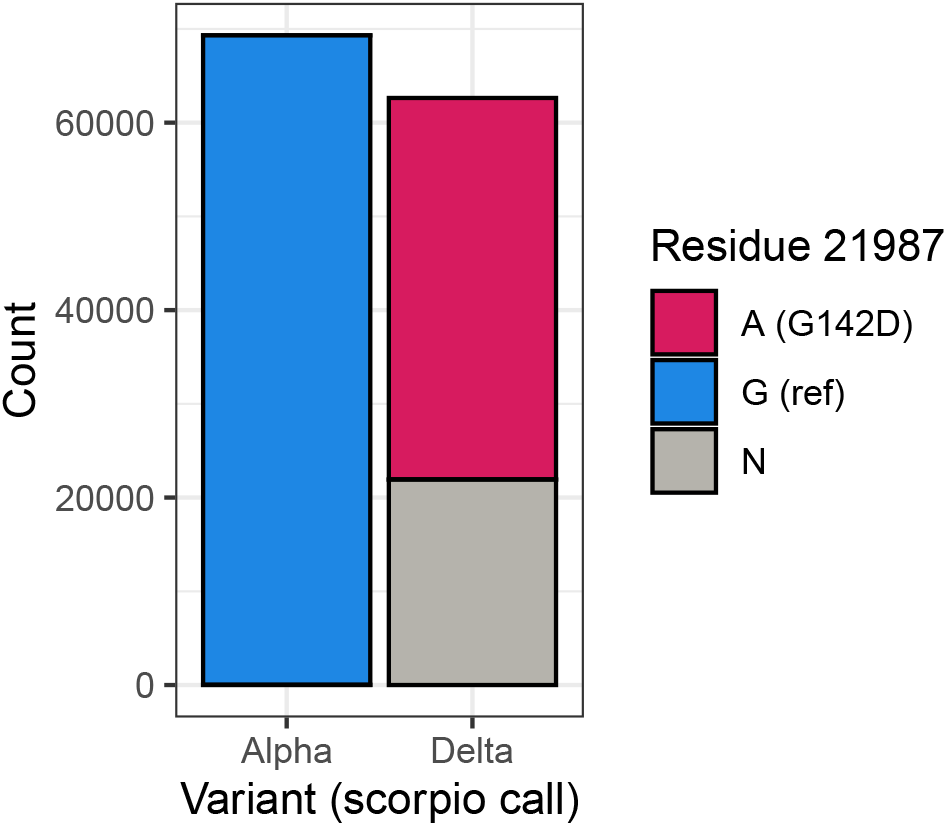
G142D is fixed in Delta, with almost all Delta sequences where the nucelotide at position 21987 is known having an A at this position. In contrast Alpha contains the reference G at this position. 35% of Delta sequences have an N at this position, indicating that the position does not have sequencing coverage.

### Missing data at Spike position 142 results from reduced coverage of ARTIC V3 amplicon 72, caused by a deletion in the binding region for its right-hand primer

To better understand how sequencing coverage of amplicon 72 affects these mutations, we examined the depth of coverage in this area for 4 sets of 10 representative genomes (Fig. 2). The top panel shows coverage of Delta sequences with position 21987 A (red) and N (grey). All these sequences show a >50-fold drop in coverage for amplicon 72, compared to 71 and 73, and for about 1/3 of sequences the coverage is so low that no consensus sequence is called, resulting in the N at this position. This phenomenon is caused by a Delta-lineage defining deletion at positions 22029– 22034, which is in the binding site for the right hand primer for amplicon 72 (found at coordinates 21904–21933, Fig. 3). The deletion removes the region to which the 5’ end of the primer would bind, reducing the binding site by five nucleotides, and *T*_*m*_ from 66 °C to 60 °C. The deletion itself is covered by amplicon 73, so it is not affected by the amplicon 72 drop-out, and is called consistently in Delta viruses.

**Fig. 2.**
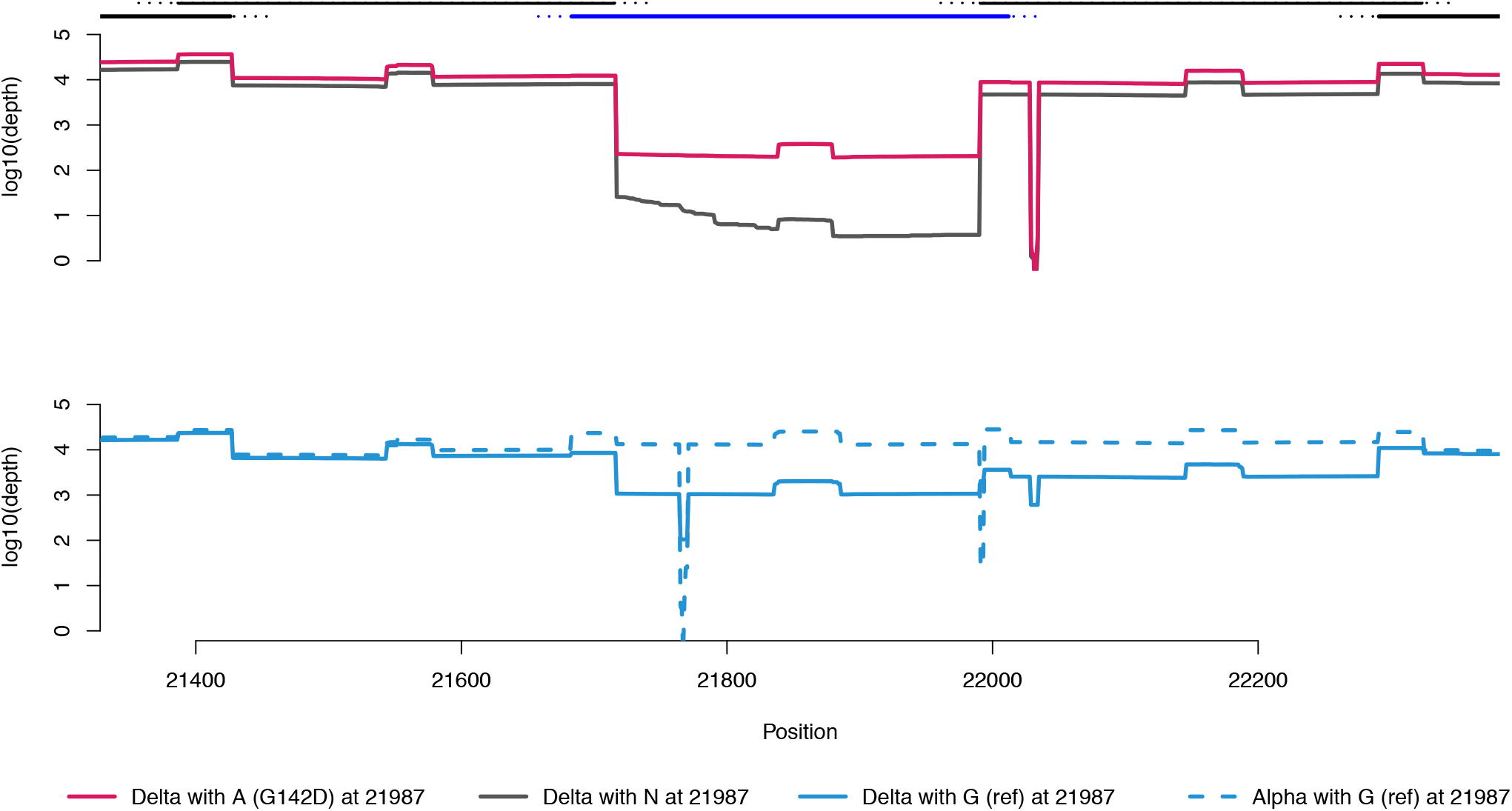
Local sequencing coverage (note log Y axis) near amplicon 72. Lines along the top show amplicons 71, 72 (blue) and 73, as well as parts of amplicons 70 and 74. Dashed portions are primers (so ought not to be present in the final sequence). In the top panel, the red line is average coverage for 10 random B.1.617.2 sequences with D at Spike position 142; grey line average coverage for 10 random B.1.617.2 sequences with N (missing data) at that position. Bumps in coverage are caused by overlap of adjacent amplicons and overlap of paired-end illumina sequence reads in the middle of each amplicon. The B.1.617.2 lineage defining deletion at 22029–22034 causes the zero coverage dip in the right-hand primer for amplicon 72. In the bottom panel, Alpha genomes (blue-dashed) have good coverage throughout, and show dips at two characteristic deletions. The blue-solid line shows coverage for the small number of sequences typed as Delta with G at Spike 142, which appear to be mixtures of Delta and Alpha.

**Fig. 3.**
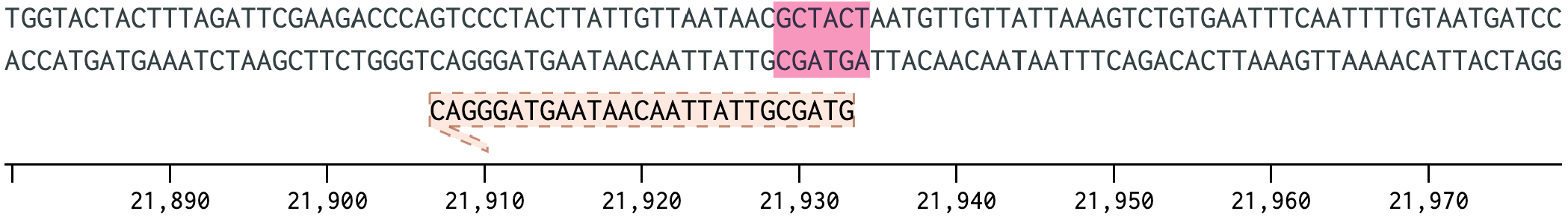
72_RIGHT primer of the ARTIC V3 scheme shown against the SARS-CoV-2 reference genome, with the lineage defining deletion at 22029–22034 in Delta highlighted.

### Remaining apparent reversions in UK data result mostly from mixed infections or contamination

Almost all sequences (>99.9%) in our dataset had an A or N at position 21987. Nevertheless, in this dataset there were 31 sequences typed as Delta (6) which had G at position 21987, and would thus be candidate “revertants”, where a second mutation, back to the reference allele, occurred at that position. We examined these in more detail. A possible explanation for this would be if the sample contained a mixture of lineages, either in the patient during a mixed infection, or from laboratory contamination. If the other lineage is present at a level too low to affect the majority of the genome but amplifies successfully for amplicon 72, then the resultant genome would appear as a mosaic, with Delta sequence everywhere apart from at amplicon 72, which would be the other lineage. Given the time period of the study the most likely candidate would be B.1.1.7 (Alpha), which has a 6 base-pair deletion at position 21764 within amplicon 72, resulting in H69/70del. We looked to see whether we could see this deletion (“-”) in the apparent revertant sequences (those with G at 21987). In the majority of cases we could (Fig. 4), suggesting that this explanation explains most of the apparent revertants in our dataset. The bottom panel of Fig. 2 shows concurrent Alpha sequences (dashed blue) have even coverage throughout this region, as well as the H69/70 deletion (and another at spike amino acid 144). The bottom solid blue line shows Delta sequences with a G at 21987, which show both intermediate coverage and mixed evidence for all three deletions (two from Alpha, one from Delta).

**Fig. 4.**
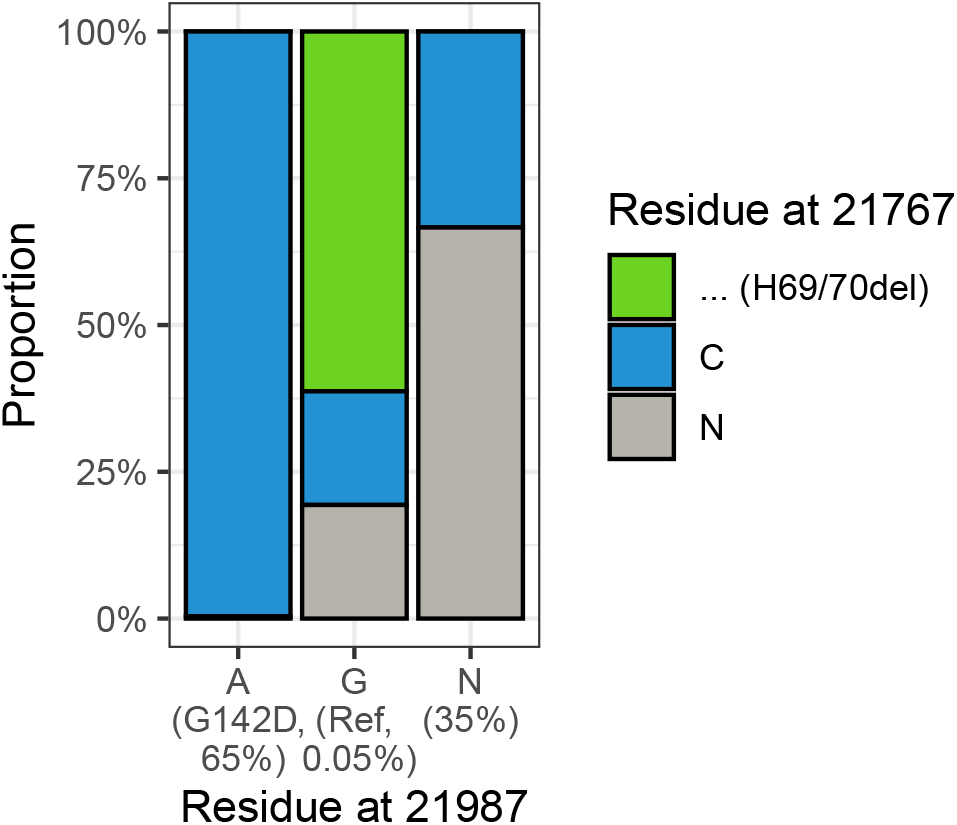
Relationship of the residue at 21987 in Delta lineage samples (representing Spike 142) with the residue at 21767 representing Spike 69/70del for the set of UK samples. Most of the small number of ‘revertants’ with G at position 21987 also have a gap at 21767, corresponding to the Spike 79/70 deletion found in B.1.1.7. This suggests that these sequences represent contamination from B.1.1.7 (either as a mixed infection or in the laboratory) that is specific to amplicon 72 because of the reduced efficiency of this amplicon in Delta samples.

### Apparent revertants in global sequencing data result mostly from untrimmed primer sequences

In a global dataset available on GISAID (7), 16% of Delta sequences have a G at postion 21987, which is a far higher rate than the tiny number of apparent Alpha contaminants in the UK data described above. Indeed, testing for the presence of H69/70del suggests only a small proportion of these can be explained by contamination or mixed infections with B.1.1.7. Residue 21987 lies within the 73_LEFT primer (21961–90) used for amplifying the amplicon immediately following amplicon 72. According to protocols, these primer sequences should be trimmed from sequencing reads prior to down-stream analysis, but if this is not performed correctly, it could lead to miscalling position 21987, since the 73_LEFT primer contains the reference sequence for this region, with a G at position 21987, and because the reduced coverage of amplicon 72 would leave fewer actual viral reads to compete with those derived from the primer.

To investigate this effect we took a random sample of GISAID genomes for which the following conditions were satisfied:

- The genome is classified as Delta
- The genome has G at position 21987
- The mapped reads that led to the consensus sequence are deposited^1^ in the Sequence Read Archive (8).

We sampled 7 such genomes, all of which were from the USA (in part because many countries do not submit to the SRA). We downloaded the raw reads that had created these genomes and examined them to look for what evidence there was for the base at position 21987 (Fig. 5).

**Fig. 5.**
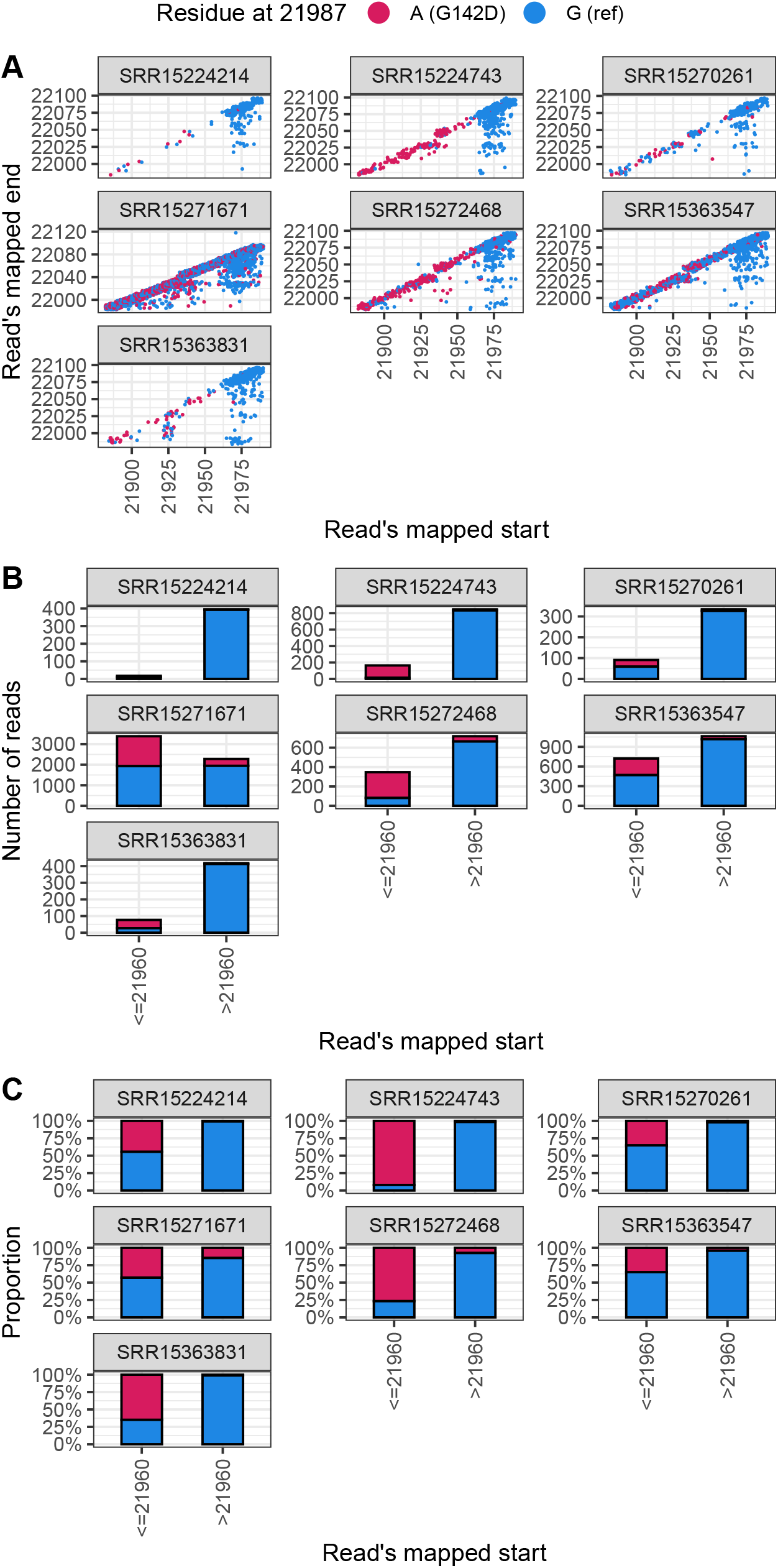
G residues at position 21987 in Delta result in large part from reads which appear to have come from the 73_LEFT primer. This plot shows 7 samples randomly selected from the SRA with a G at position 21987 of the consensus sequence. In all cases there is substantial evidence for A at this position. Specifically reads which must have come from amplicon 72 (indicated as <= 29160) show substantial evidence for A, whereas reads likely to have come from the 73_LEFT primer sequence (indicated as *>* 29160) are overwhelmingly G. (Panel A features some jitter to reduce overplotting.)

In all cases there were a substantial number of reads with an A at position 21987. Plotting reads according their starting point in the genome, to simulate the effect of primer clipping, suggested that in all the cases examined clipped reads provided reasonable evidence for the possibility of A at at position 21987, with 3 cases where A was the significant majority residue in clipped reads.

However not all reads beginning prior to 29160 had an A at position 21987. An explanation has very recently been provided by others (9), who show that the pools containing the primers for ARTIC amplicons 71 and 73 can result in nonspecific amplification of a hybrid amplicon that also incorporates amplicon 72 sequence (as well as the 73_LEFT primer sequence). This would also mean that even when clipping was carried out (which our analysis suggests it sometimes is not), the primer sequence could be presented in an unexpected context in which it would not be removed. This effect is particularly difficult to diagnose and correct in the case of short read sequencing.

### T95I is subject to the same dropout effect, but is also genuinely limited to certain Delta sublineages

We will briefly discuss T95I, which was suggested to exhibit similar dynamics to G142D (3). T95I corresponds to a mutation at nucleotide 21846 from C to T. This position is also found within amplicon 72 and so is expected to exhibit similar artifacts to those described above.

T95I differs from G142D in that this mutation is *not* fixed within Delta. Rather there is genuine underlying biological variation of genotypes, over which the technical effects described above are layered. This can be seen in Fig. 6, which shows that T95I is fixed in the AY.4 sublineage of Delta, but absent from any of the other designated sublineages. As anticipated, a substantial amount of sequences from all Delta sublineages have N at this position, reflecting the reduced efficiency of amplification at amplicon 72. As previously, this technical effect can explain the relationship between detection of T95I and higher viral loads reported in Shen et al. (3).

**Fig. 6.**
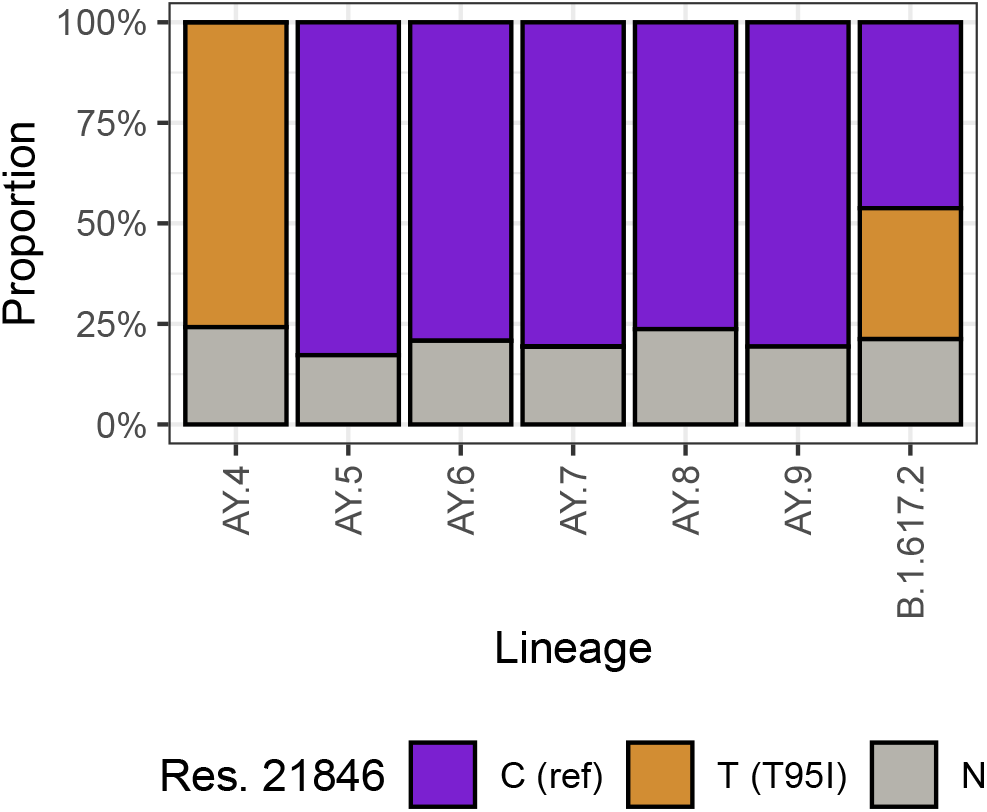
Distribution of nucleotides at 21846 (nucleotide T encodes T95I) for different sublineages within Delta. T95I is fixed in AY.4, but absent from other currently designated Delta sublineages.

### An increased likelihood of correctly calling position 142 for high viral load samples creates a spurious correlation between *C*_*t*_ values and genotype

These observations provide a clear hypothesis that could explain the observation of a correlation between viral load and the presence of the G142D mutation reported in Shen et al. (3). For samples with low viral load (high *C*_*t*_ values), one might expect that the reduced amplification efficiency for amplicon 72 would be more likely to lead to an N at position 21987 than for samples with higher viral loads. To formally test this we plotted the residue at 21987 by *C*_*t*_ value, and indeed found that though at average *C*_*t*_ values the residue was able to be called correctly as A, as *C*_*t*_ value approached 30 almost all calls became Ns ^2^ (Fig. 7). It is therefore likely that the observed relationship between genotype and *C*_*t*_ value is caused not by the genotype affecting the viral load, but instead the reverse, with the amount of viral material in the sample affecting the ability to detect the genotype.

**Fig. 7.**
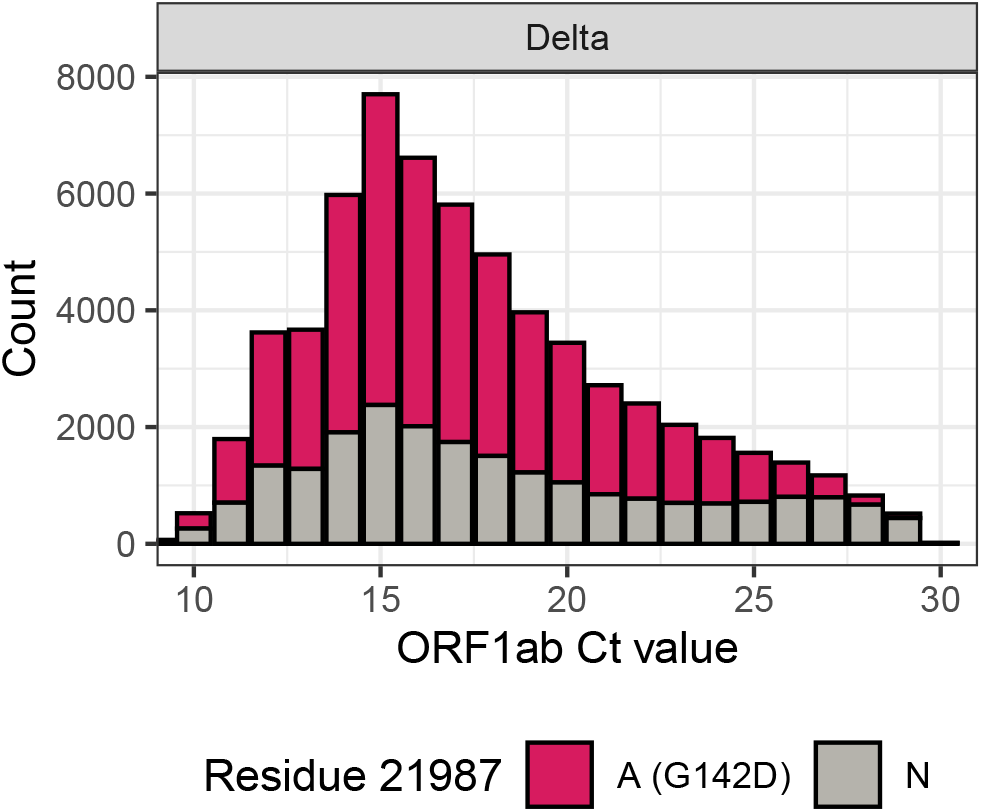
Relationship of Ct value and residue at position 21987 for COG-UK Delta samples until 30 June 2021

## Discussion

We have shown the apparent diversity within Delta at Spike 142 and immediately adjacent regions reflects reduced amplification of a sequencing region rather than underlying biology. G142D is fixed in Delta, with essentially all apparent back mutations being artifacts. Similarly, T95I is fixed in AY.4, and any apparent reversions in that clade are artifactual. This suggests there is no current basis to expect a biological causative relationship between the presence of G142D or T95I and viral load.

For most analyses of Delta sequences it is likely to be advantageous to mask out the entire amplicon 72 sequence to avoid being misled by the various technical effects at these sites. In general when analysing consensus sequences processed with unknown pipelines, masking out ARTIC V3 primers is often likely to be desirable – for example D950N is another Delta mutation found within an ARTIC V3 primer site which exhibits spurious apparent reversions in global Delta sequences. Assumptions that the failure to detect a given mutation implies the absence of that mutation are prone to mistakes – it is important to consider the alternative that we lack information about a particular site.

Regular monitoring of amplicon coverage may assist in early detection of mutations at primer binding sites, and the artifacts that these may cause. Trimming primer sequences is an essential part of any pipeline for generation of consensus sequences, but may not be being performed robustly for a significant proportion of current global sequences. Importantly, open deposition of raw reads allowed us to examine these effects, and could even allow large-scale correction of erroneous sequences if suitable infrastructure were developed. The non-specific amplification of amplicon 72 sequence noted by Cerutti et al. (9) further complicates these effects.

The ARTIC Network has released a V4 of the amplicon scheme (10), which resolves these issues (11), and we would encourage researchers to move to it to allow detection across the genome given that almost all sequenced genomes are currently Delta.

## Data Availability

We analyse three datasets here:
Viral genomes sequenced by the Sanger Institute at part of the COG-UK Consortium. These can be accessed at https://cog-uk.s3.climb.ac.uk/phylogenetics/latest/cog_all.fasta, or as an alignment at https://cog-uk.s3.climb.ac.uk/phylogenetics/latest/cog_alignment.fasta. Metadata, including lineage calls, is at https://cog-uk.s3.climb.ac.uk/phylogenetics/latest/cog_metadata.csv. Global SARS-CoV-2 genomes available at https://gisaid.org. An alignment is also available there within the Downloads section, with a separate metadata file, including lineage calls. Raw reads from the Sequence Read Archive were accessed from https://www.ncbi.nlm.nih.gov/sra Fully deidentified Ct-genotype mappings, as well as the other COG-UK data analysed are available at https://github.com/theosanderson/amplicon_72.

## Acknowledgements

We thank the thousands of labs around the world who have deposited SARS-CoV-2 sequence data to public databases like GISAID and the SRA, as well as the Wellcome Sanger Institute Covid-19 Surveillance Team and COVID-19 Genomics UK (COG-UK) consortium. We also acknowledge @babarlelephant’s early insights into these issues.

## Methods and reproducibility

Further information on the methods is available in the *Supplementary methods* file. Our code can be accessed at https://github.com/theosanderson/amplicon_72.

## Footnotes

1 We used https://github.com/nextstrain/ncov-ingest/blob/05b3b36d8264f017b1a931a5427903793cbef802/source-data/accessions.tsv to convert GISAID IDs to GenBank IDs, which we then scanned for SRA accessions

2 A trend towards more N calls at extremely low *C*_*t*_ values (high viral loads, *C*_*t*_ ≈ 10) was also seen, which may reflect very large amounts of DNA template titrating out primers, leading to reduced coverage at this amplicon.

## Bibliography

1. Josh Quick, Nick Loman, and ARTIC Network. Artic.network, 2020. URL https://artic.network/resources/ncov/ncov-amplicon-v3.pdf.

2. J. R. Tyson, P. James, D. Stoddart, N. Sparks, A. Wickenhagen, G. Hall, J. H. Choi, H. Lapointe, K. Kamelian, A. D. Smith, N. Prystajecky, I. Goodfellow, S. J. Wilson, R. Harrigan, T. P. Snutch, N. J. Loman, and J. Quick. Improvements to the ARTIC multiplex PCR method for SARS-CoV-2 genome sequencing using nanopore. bioRxiv, Sep 2020.

3. Lishuang Shen, Timothy J. Triche, Jennifer Dien Bard, Jaclyn A. Biegel, Alexander R. Judkins, and Xiaowu Gai. Spike protein ntd mutation G142D in sars-cov-2 delta voc lineages is associated with frequent back mutations, increased viral loads, and immune evasion. medRxiv, 2021. doi: 10.1101/2021.09.12.21263475. URL https://www.medrxiv.org/content/early/2021/09/15/2021.09.12.21263475.

4. S. M. Nicholls, R. Poplawski, M. J. Bull, A. Underwood, M. Chapman, K. Abu-Dahab, B. Taylor, R. M. Colquhoun, W. P. M. Rowe, B. Jackson, V. Hill, Á. O’Toole, S. Rey, J. Southgate, R. Amato, R. Livett, S. Gonçalves, E. M. Harrison, S. J. Peacock, D. M. Aanensen, A. Rambaut, T. R. Connor, and N. J. Loman. CLIMB-COVID: continuous integration supporting decentralised sequencing for SARS-CoV-2 genomic surveillance. Genome Biol, 22(1):196, 07 2021.

5. Harald S. Vöhringer, Theo Sanderson, Matthew Sinnott, Nicola De Maio, Thuy Nguyen, Richard Goater, Frank Schwach, Ian Harrison, Joel Hellewell, Cristina Ariani, Sonia Gonçalves, David Jackson, Ian Johnston, Alexander W. Jung, Callum Saint, John Sillitoe, Maria Suciu, Nick Goldman, Jasmina Panovska-Griffiths, The Wellcome Sanger Institute Covid-19 Surveillance Team, The COVID-19 Genomics UK (COG-UK) Consortium, Ewan Birney, Erik Volz, Sebastian Funk, Dominic Kwiatkowski, Meera Chand, Inigo Martincorena, Jeffrey C. Barrett, and Moritz Gerstung. Genomic reconstruction of the sars-cov-2 epidemic in england. medRxiv, 2021. doi: 10.1101/2021.05.22.21257633. URL https://www.medrxiv.org/content/early/2021/08/17/2021.05.22.21257633.

6. Á. O’Toole, E. Scher, A. Underwood, B. Jackson, V. Hill, J. T. McCrone, R. Colquhoun, C. Ruis, K. Abu-Dahab, B. Taylor, C. Yeats, L. du Plessis, D. Maloney, N. Medd, S. W. Attwood, D. M. Aanensen, E. C. Holmes, O. G. Pybus, and A. Rambaut. Assignment of epidemiological lineages in an emerging pandemic using the pangolin tool. Virus Evol, 7(2): veab064, 2021.

7. S. Elbe and G. Buckland-Merrett. Data, disease and diplomacy: GISAID’s innovative contribution to global health. Glob Chall, 1(1):33–46, Jan 2017.

8. R. Leinonen, H. Sugawara, and M. Shumway. The sequence read archive. Nucleic Acids Res, 39(Database issue):19–21, Jan 2011.

9. Lorenzo Cerutti. Missing g21987a mutation in sars-cov-2 delta variants due to non-specific amplification by artic v3 primers. Virological.org, 2021. URL https://virological.org/t/missing-g21987a-mutation-in-sars-cov-2-delta-variants-due-to-non-specific-amplification-by-artic-v3-primers/764.

10. Josh Quick and ARTIC Network. Artic.network, 2021. URL https://community.artic.network/t/sars-cov-2-version-4-scheme-release/312.

11. James J. Davis, S. Wesley Long, Paul A. Christensen, Randall J. Olsen, Robert Olson, Maulik Shukla, Sishir Subedi, Rick Stevens, and James M. Musser. Analysis of the artic version 3 and version 4 sars-cov-2 primers and their impact on the detection of the g142d amino acid substitution in the spike protein. bioRxiv, 2021. doi: 10.1101/2021.09.27.461949. URL https://www.biorxiv.org/content/early/2021/10/08/2021.09.27.461949.

