## Supplementary methods for "Variation at Spike position 142 in SARS-CoV-2 Delta genomes is a technical artifact caused by dropout of a sequencing amplicon"

### Supplementary materials and methods

We have made a git repository (https://github.com/theosanderson/amplicon_72) available containing our code, and an R Notebook reproducing COG-UK figures in this paper.
COG-UK data is publicly available, allowing complete reproduction of our results. Accessing GISAID data will require the user to have an account, or to apply for one at EpiCov.org, and downloading the data.

#### Codon2Nucleotide

Codon2Nucleotide is a React app, which is open-source. We have made the source code available at https://github.com/theosanderson/Codon2Nucleotide. Codon2Nucleotide can easily be adapted to other organisms with relatively small genomes. It does not currently provide facilities for exons.

#### Residue extraction

We used a simple Python script (see repo) to extract residues at certain locations from COG-UK and GISAID multiple sequence alignments, created from consensus sequences aligned to the Wuhan Hu-1 reference sequence.

#### Coverage plots

Coverage was calculated with `samtools depth`, from 10 random genomes satisfying each condition. BAM files are available for all COG-UK genomes sequenced at the Sanger Institute through the ENA or SRA (https://www.ncbi.nlm.nih.gov/sra).

#### Analysis of individual genome reads

Firstly we mapped GISAID genomes to their references in the SRA, by firstly connecting each GISAID to its GenBank entry (where available) using the file available at https://github.com/nextstrain/ncov-ingest/blob/05b3b36d8264f017b1a931a5427903793cbef802/source-data/accessions.tsv. We then queried GenBank for corresponding SRA accessions using the get$_{s}ra_{i}ds.pycustomPythonscriptthatcanbefoundintherepo.$
We downloaded 7 random genomes, and used the `create_pileup.py` script in the repo to create a pile-up with the `pysam` library and extract the residue at each position, as well as the start and end positions of the read.
